# Negative nasopharyngeal SARS-CoV-2 PCR conversion in Response to different therapeutic interventions

**DOI:** 10.1101/2020.05.08.20095679

**Authors:** Mohammed Hassan Shabrawishi, Abdallah Y Naser, Hassan Alwafi, Ahmad Mansoor Aldobyany, Abdelfattah Ahmed Touman

## Abstract

**BACKGROUND:** Despite lack of convincing evidence of the efficacy of hydroxychloroquine, it has been suggested to be used for the treatment of SARS-CoV-2 to accelerate the negative virus conversion. We aimed to explore the association between negative nasopharyngeal SARS-CoV-2 PCR clearance and different therapeutic interventions.

**METHODOLOGY:** This was a retrospective cohort study of 93 patients who were admitted to medical ward with a PCR confirmed diagnosis of COVID-19 and met the inclusion criteria in a tertiary hospital in Mecca, Saudi Arabia. There were three interventional subgroups (group A (n=45): who received antimalarial drug only classified as (A1), combined with azithromycin (A2) or combined with antiviral drugs (A3)), and one supportive care group (group B) (n=48). The primary and secondary endpoints of the study were achieving negative SARS-CoV-2 nasopharyngeal PCR sample within five days or less from the start of the intervention and 12 days or less from the diagnose, respectively.

**RESULTS:** The mean age of the patients was 43.9 years (SD:15.9). A median time of 3.00 days (IQR:2.00-6.50) needed from the time of starting the intervention/supportive care to the first negative PCR sample. There was no statistically significant difference neither between the percentage of patients in the interventional group and the supportive care group who achieved the primary or the secondary endpoint, nor in the median time needed to achieve the first negative PCR sample (p>0.05).

**CONCLUSION:** Prescribing antimalarial medications was not shown to shorten the disease course nor to accelerate the negative PCR conversion rate.

## INTRODUCTION

In December 2019, a novel corona virus named SARS-CoV-2 emerged in china and spread worldwide to be declared by WHO as a pandemic in 12 March 2020 (1). Patients with COVID-19 present with fever, dry cough and shortness of breath, however, some are asymptomatic. The majority of cases have favourable outcomes, nonetheless, older patients and patients with comorbidities may have worse outcomes (2).

SARS-CoV-2 spread mainly through respiratory droplet and close contact. Nevertheless, studies have shown that asymptomatic carriers can be contagious as well (3, 4). Health care systems in many countries falls under tremendous pressure of an increasing number of confirmed cases, and many heath care authorities recommended two negative nasopharyngeal polymerase chain reaction (PCR) result 24 hours apart before discontinuation of hospital isolation (5, 6). To this date, there are no therapeutic options approved by the U.S. Food and Drug Administration (FDA) for the prevention or the treatment of COVID-19. Current management practices consist of infection prevention and supportive care, such as oxygen supplementation and mechanical ventilation if needed (7). Many studies have been conducted to identify effective treatment in order to cure symptomatic patients and to limit the transmission to the community. Different medications were proposed to be candidate for the treatment of COVID-19, some of these options focused on the use of old antiviral medications and testing its effectiveness against COVID-19 (8, 9). There are contradictory findings against the effectiveness of antimalarial agents such as hydroxychloroquine (HCQ) and chloroquine on COVID-19. Many studies have demonstrated their effectiveness in inhibiting SARS-CoV-2 (10-12). In a recent clinical trial, HCQ reported to cause significant reduction in viral carriage at the day number six post inclusion, with around 70% of patients having negative nasopharyngeal PCR sample, compared to untreated patients (12.5%) (13). On the other hand, a Chinese study reported no significant differences between patients who received HCQ and the control group regarding pharyngeal carriage of viral RNA at day seven (14). Despite that there are more than 80 trials registered to investigate the effectiveness of these antimalarial agents against COVID-19 as a monotherapy or combination with other medications, these trials characterized by having poor methodological aspects and reporting (15). In addition, the use HCQ might expose some patients to different life-threatening consequences such as cutaneous adverse reactions, fulminant hepatic failure, and ventricular arrhythmias (especially when prescribed with azithromycin) (16-19).

A recent Chinese study focused on the duration of viral shedding and reported a median duration between 20.0 days and up to 37.0 days among survivors (20). Another single centre French study explored the use of combination of oral HCQ sulfate with azithromycin, and reported that this combination was able to negatively convert the nasopharyngeal viral load as tested by PCR in all studied cases in day 12 (21). Accelerating the negative virus conversion allow for earlier discharge from the hospital and/or designated isolation facilities and facilitate a more efficient utilization of the health care bed capacity. In this study, we aimed to study the association between negative nasopharyngeal SARS-CoV-2 PCR conversion and different therapeutic interventions (HCQ monotherapy, as combination with macrolide, or as combination with antiviral with or without azithromycin).

## METHODOLOGY

### Setting

This study was conducted between 7th March until 15th April 2020, at the inpatient medical ward of Al-Noor Specialist hospital, a tertiary public hospital in Mecca, Kingdom of Saudi Arabia.

### Study design

A retrospective cohort study that included 145 patients who were symptomatic and have PCR confirmed diagnosis of novel corona virus disease (COVID-19). Al-Noor Specialist hospital is a designated referral center for confirmed COVID-19 cases in Mecca province, KSA. The majority of COVID-19 cases were screened in other facilities and referred to the hospital after being confirmed positive for SARS-COV-2. All PCR samples is being sent and processed in a regional laboratory. We choose the negative conversion rate of SARS-CoV-2 nasopharyngeal PCR at day 5 and negative conversion at day 12 from the first positive sample as our primary and secondary endpoints as it was shown to adequate measure of treatment effectiveness in Philippe Gautreta et al. studies (13, 21). At the time we conducted this study the Saudi ministry of health (MOH) recommended interventions for confirmed cases only, thus all patients were receiving best supporting care only until positive PCR result confirmed. Furthermore, MOH guideline suggest (optional) to start antiviral and or HCQ after approval of an infectious disease consultant.

### Study population

We included all PCR confirmed cases of SARS-COV-2 admitted to the medical ward at Al-Noor Specialist hospital. Hospital protocol of retesting of symptomatic patients follow the Saudi Center for Disease Prevention and Control recommendations, that recommends re-testing when a patient is clinically recovered, and to be repeated every 72 hours if the result remain positive (6). Our exclusion criteria were; 1) patients less than 12 years of age, 2) cases that were directly admitted to the intensive care unit (ICU), 3) patients who develop critically severe disease and were shifted to the ICU while still showing a positive nasopharyngeal PCR result for SARS-COV-2, and 4) clinically stable patients who were transferred to the ministry of health (MOH) designated isolation facilities while still having positive PCR results. Exclusion criteria two and three were defined because the retesting protocol at the ICU is performed irregularly and with long interval periods

### Outcomes

The primary endpoint of the study is achieving negative SARS-CoV-2 nasopharyngeal PCR within five days or less from the start of the intervention. Secondary endpoint was achieving negative sample within 12 days or less from the first positive PCR result.

### Intervention and control groups

Patients were categorised into two main groups; group A that includes patients who received any active interventions. We defined active interventions as patients who received any of the following medications: chloroquine, HCQ, Ribavirin and/or lopinavir and ritonavir. On the other hand, group B was defined as the best supportive care group. Patients who did not receive any dose of the active interventional drugs were categorised into group B.

Group A, were subsequently sub-grouped into A1 A2, and A3. Subgroup A1 includes patients who received any dose of chloroquine or HCQ without any dose of azithromycin antibiotic or antivirals (Ribavirin and/or lopinavir and ritonavir). Subgroup A2 includes patients received any dose of chloroquine or HCQ and any dose of azithromycin not necessary simultaneously. Subgroup A3 (multiple interventions subgroups) includes patients received any dose of chloroquine or HCQ and any dose of any antiviral drugs (Ribavirin and/or lopinavir and ritonavir) with or without azithromycin. **Figure 1** below highlights the flow chart for the study inclusion.

### Definitions

Severity of the disease were defined as the following; 1) mild disease was defined as patient with upper respiratory tract symptoms (as rhinorrhea, sore throat, headache, myalgia, body pain, low grad fever and or dry cough) with absent of clinical or radiological finding of pneumonia; 2) moderate disease was defined as symptomatic patient with either clinical or radiological sign of pneumonia; 3) severe disease was defined as confirmed COVID-19 pneumonia with any of the following: respiratory rate ≥30/min, blood oxygen saturation ≤93% at rest, PaO2/FiO2 ratio <300, lung infiltration >50% of the lung field, and 4) critically severe disease defined with any of the following: respiratory failure required invasive mechanical ventilation, shock or organ failure require admission to the intensive care unit.

#### Negative conversion

We used the first negative nasopharyngeal PCR test to define the negative conversion.

#### Time to negativity definition

Days to negative PCR clearance was calculated from the first positive sample to the first negative sample. Days between starting the medical management to the achievement of PCR negative clearance was calculated from the first dose of HCQ or chloroquine in subgroup A1 and A2. In subgroup A3, it was calculated from the first dose of any given intervention whether the antimalarial drug or the antiviral medications (whichever first). In regard the group B, it was calculated starting from the date of admission to the date of negative PCR clearance. As a referral center, the admission date lag behind the first positive PCR results.

### 2.6 Ethical approval

The study protocol was reviewed and approved the institutional ethics board at the Ministry of Health in Saudi Arabia (No. H-02-K-076-0420-286). All the recruited subjects provided written consents.

### 2.7 Statistical analysis

Data were analysed using the SPSS software, version 25. The descriptive analysis was reported as mean (± standard deviation [SD]) for normally distributed quantitative variables and as median (interquartile range [IQR]) for non-normally distributed quantitative variables. Kolmogorov Simonov, Shapiro Wilk, and histogram tests were used to check the normality of the data. Categorical data were reported as percentages and frequencies. The Mann-Whitney U test/Kruskal-Wallis test was used to compare the median days to achieve PCR clearance between different demographic groups. In addition, logistic regression analysis was applied to identify factors associated with PCR clearance. A confidence interval of 95% (p < 0.05) was applied to represent the statistical significance of the results and the level of significance was assigned as 5%.

## RESULT

### Baseline characteristics

A total of 93 out of 145 patients (64.1%) who have met the inclusion criteria were included in this study. From the 52 patients who were excluded, 43 were clinically stable and were transferred to the MOH designated isolation facilities while still having positive PCR results. Nine patients were excluded as they were critically ill and shifted to the ICU.

A total of 45 patients (48.4%) formed the interventional group (Group A), while 48 patients met criteria of best supportive care group. From the interventional group A; 15, 25, and 5 patients met criteria for subgroups A1, A2, and A3 respectively. The majority of the patients in the interventional group were males 27 (60%), contrary, females were the majority in group B with 26 patients (54.2%). Group A had significantly more severe disease with 9 patients (20.0%) presented with severe illness compared to one patient (2.1%) in group B (p<0.000). The majority of patients received best supportive care had mild disease (85.4%, n= 41). Moderately sever illness were 44.4% and 12.5% in group A and B, respectively. **Table 1** below presents patients baselines demographics.

### Effect of interventions

As it is highlighted in **Figure 1**, Subgroup A1 patients received HCQ and subgroup A2 patients received combination of HCQ and macrolide. All the patients (n=5) in subgroup A3 received azithromycin and HCQ. Three patients received lopinavir/ritonavir combined with ribavirin, while, two have received it without ribavirin.

All patients in this study needed a median time of 6.00 days (4.50 – 9.00) from first positive to the first negative PCR sample, and a median time of 3.00 days (2.00 – 6.50) from the time of starting the intervention to the first negative PCR sample. Around 71.0% (n= 66) and 81.7% (n= 76) of the patients achieved the primary and secondary endpoint, respectively. There was no statistically significant difference between the percentage of patients in the interventional group (group A) and the supportive care group (group B) who achieved the primary or the secondary endpoint (p>0.05). In group A 73.3% (n= 33) achieved the primary endpoint and 84.4% (n= 38) achieved the secondary endpoint. Smaller percentage of patients 68.8 (n= 33) and 79.2% (n= 38) achieved the primary and secondary endpoints in group B. There was no statistically significant difference in the median time to negative conversion from the first positive to the first negative PCR sample or from the time of starting the intervention between the two groups (p>0.05) **Table 2**.

### Patients characteristics and PCR clearance time

**Table 3** details the median time needed to achieve the first negative PCR sample from the first positive and from intervention stratified by patients’ demographics and treatment groups. There was statistically significant difference in the median time from first positive and intervention to first negative PCR sample between elderly patients (aged 45 years and above) and younger (p<0.01). In addition, median time from intervention to first negative PCR sample significantly differed by disease severity (p<0.05). On the other hand, there was no statistically significant difference between the intervention group (group A) and the supportive care group (group B) in term of the median time from first positive and intervention to first negative PCR sample (p>0.05).

Binary logistic regression analysis showed that males were 3.9 times more likely to achieve primary endpoint and achieve negative PCR sample within 5 days compared to females (OR: 3.90 (95%CI 1.49 – 10.22). In addition, males were 4.7 times more likely to achieve secondary endpoint and achieve negative PCR sample within 12 days compared to females (OR: 4.71 (95%CI 1.41 – 15.83). On the other hand, age was negatively associated with achieving the primary and secondary endpoints and elderly patients aged 45 years and above were less likely to achieve them by around 77.0% (OR: 0.23 (95%CI 0.09 – 0.59) and 93.0% (OR: 0.07 (95%CI 0.02 – 0.35), respectively.

Using multiple logistic regression, we applied two models: the first one to explore the effect of the intervention subgroups (A1, A2, and A3) compared to supportive care group (group B) on achieving the primary endpoint (achieving negative PCR sample within 5 days or less) adjusting for age, gender, and disease severity, and the second model to explore the effect on achieving the secondary endpoint (achieving negative PCR sample within 12 days or less). The first model did not find any statistically significant difference between any intervention subgroup and the supportive care group in achieving the primary endpoint (p>0.05). The second model found negative association between the intervention subgroup A3 and achieving the secondary endpoint. Patients in subgroup A3 were around 97.0% less likely to achieve the secondary endpoint (OR: 0.033 (95%CI 0.001 - 0.863). However, the association was very week (p= 0.040).

## DISCUSSION

This was a retrospective cohort study of patients who were admitted with a PCR confirmed diagnosis of COVID-19. We investigated the association between negative nasopharyngeal SARS-CoV-2 PCR clearance and different therapeutic interventions. This study found that the use of HCQ for the treatment of COVID-19 whether as monotherapy or in combination therapy was not significantly associated with better negative PCR clearance or shorter time compared to supportive care. Males were f to 5 times more likely to achieve negative PCR clearance compared to females within 5 days and 12 days, respectively. In addition, age was negatively associated with achieving negative PCR clearance within 5 days and 12 days.

To date there is no proven effective therapy for SRAS-COV2, however, HCQ was adopted as an optional therapy after an encouraging initial in vitro result (12). Despite lack of convincing evidence of its efficacy, HCQ has been suggested to be used by different medical regularity authorities based on small, non-randomized promising clinical studies (13, 21, 22).

The findings of our study aligns with previous studies that demonstrated no superior value for the administration of HCQ in treating COVID-19 (14, 15). The baseline demographic characteristics in our study showed comparable age across the two groups. However, group A (treated using HCQ, with or without azithromycin and additional antiviral therapy) was more likely to be males (60.0%). Moreover, group A specially subgroup A3 had more severe disease. There was no statistically significant difference in the median time required to the first negative PCR sample between group A and group B (p>0.05). Compared to Philippe Gautreta et al. study where the number of contagious patients dropped to zero on day12; 18.3% of our patients who received interventions remained positive after the 12^th^ day. Despite that there was a difference in the baseline severity of the cases between the patients who received pharmacological therapy (using HCQ as monotherapy or combination therapy) (group A) and the patients who received supportive care (group B), we did not found any statistically significant difference in the rate of achieving negative PCR clearance or in the time needed to achieve it (p>0.05). There was no statistically significant difference between percentage of patients in group A who achieved primary and secondary endpoint (73.3% and 84.4%) compared to group B (68.8 and 79.2%) (p>0.05).

Our study showed that the interventional group did not have shorter disease course nor faster negative conversion of the nasopharyngeal swab result for SARS-COV-2. We did not study the clinical usefulness of these interventions in term of clinical improvement such as attenuation of the disease severity or mortality reduction. However, we are aware of the negative study that showed no reduction in mechanical ventilation risk in patients hospitalized with Covid-19 and received HCQ, either with or without azithromycin (23).

### Strengths and limitations

This study has several strengths. To our knowledge, this was the first study in the Middle East region to explore the association between the use of HCQ as monotherapy or combination therapy and the odds of achieving negative PCR sample in COVID-19 patients. As the data collection center was the designated regional COVID-19 center, patients were diagnosed at different locations and then transferred to our center thus our study cohort is less susceptible to selection biases of single-center studies. However, our study has some limitations. The study design was a retrospective single referral center. Thus, it inherent all retrospective analyses limitation such as non-randomization of treatments. In addition, the small sample size in our study specifically for the subgroups in the intervention group (group A) might have limited our ability to explore statistically significant difference and had led to a wide confidence intervals. Despite that we conducted multiple logistic regression to adjust for the severity of the disease and the age, patients could still have confounders that we were not able to measure and might influenced our findings.

## CONCLUSION

The study showed no significant different in time to the negative PCR clearance between patients received HCQ whether alone or combined with azithromycin and/or antivirus drugs compared with patients treated with the best supportive care. Prescribing antimalarial medications was not shown to shorten the disease course nor to accelerate the negative PCR conversion rate or the hospital discharge.

## Data Availability

The data that support the findings of this study are available from the corresponding author upon reasonable request.

## AUTHORS CONTRIBUTIONS

Shabrawishi, Naser, Touman, and Alwafi had full access to all the data in the study and take responsibility for the integrity of the data and the accuracy of the data analysis. Shabrawishi, and Touman had the original idea for this study. Shabrawishi, Naser, Touman, and Alwafi contributed to the design of the study. Shabrawishi and Touman contributed to the data collection. Naser contributed to the statistical analysis. Shabrawishi, Naser, Touman, and Alwafi wrote the first draft. All the authors contributed to interpretation and edited the draft report.

---

**Tables legends**

Table 1: Patients baseline characteristics.

Table 2: Median time and percentage of patients achieved the primary and secondary outcomes.

Table 3: Patients characteristics and PCR clearance time.

**Figures legends**

Figure 1: Study flow chart

**Table 4:** Patients baseline characteristics.

| Variable | Frequency (%) |
| --- | --- |
| **Age [mean] (years; SD)** | 43.9 years (SD: 15.9) |
| **Group A Age (years; SD)** |  |
| Subgroup A1 Age (years; SD) | 42.9 years (SD: 17.2) |
| Subgroup A2 Age (years; SD) | 46.4 years (SD: 16.4) |
| Subgroup A3 Age (years; SD) | 41.4 years (SD: 18.3) |
| **Group B Age (years; SD)** | 43.2 years (SD: 15.4) |
| **Gender** |  |
| Overall - Male No. (%) | 49 (52.7) |
| GROUP A- Male No. (%) | 27 (60.0) |
| GROUP B- Male No. (%) | 22 (45.8) |
| **Severity for all patients No. (%)** |  |
| Mild | 57 (61.3) |
| Moderate | 26 (28.0) |
| Severe | 10 (10.8) |
| **Severity group A No. (%)** |  |
| Mild | 16 (35.6) |
| Moderate | 20 (44.4) |
| Severe | 9 (20.0) |
| **Severity group B No. (%)** |  |
| Mild | 41 (85.4) |
| Moderate | 6 (12.5) |
| Severe | 1 (2.1) |

SD: standard deviation; No: frequency

**Table 5:** Median time and percentage of patients achieved the primary and secondary outcomes.

| Variable | All patients | Group A | Group B | P-value |
| --- | --- | --- | --- | --- |
| Median time from first positive to first negative PCR sample for all patients (IQR) | 6.00<br>(4.50 – 9.00) | 6.00<br>(5.00 – 9.00) | 6.00<br>(4.00 – 9.75) | 0.574 |
| Median time from intervention to first negative PCR sample for all patients (IQR) | 3.00<br>(2.00 – 6.50) | 3.00<br>(2.00 – 6.50) | 3.00<br>(2.00 – 6.75) | 0.895 |
| Patients achieved primary endpoint and had negative PCR sample within 5 days or less No. (%) | 66 (71.0) | 33 (73.3) | 33<br>(68.8) | 0.655 |
| Patients achieved secondary endpoint and had negative PCR sample within 12 days or less No. (%) | 76 (81.7) | 38 (84.4) | 38 (79.2) | 0.597 |

**Table 6:** Patients characteristics and PCR clearance time.

| Variable | Median time from intervention to first negative sample (IQR) | P-value | Median time from first positive to first negative sample (IQR) | P-value |
| --- | --- | --- | --- | --- |
| Age |  |  |  |  |
| Below 45 years | 3.00 (1.00 – 4.00) | 0.005** | 6.00 (4.00 – 7.00) | 0.003** |
| 45 years and above | 4.50 (2.00 – 11.25) |  | 8.00 (5.00 – 14.00) |  |
| Gender |  |  |  |  |
| Male | 3.00 (2.00 – 5.00) | 0.172 | 6.00 (4.00 – 8.00) | 0.095 |
| Female | 3.50 (1.00 – 9.00) |  | 7.00 (5.00 – 13.00) |  |
| Severity |  |  |  |  |
| Mild | 3.00 (1.00 - 5.50) | 0.039* | 6.00 (4.00 – 9.00) | 0.103 |
| Moderate | 4.00 (2.00 – 6.25) |  | 6.50 (5.00 – 9.00) |  |
| Severe | 6.50 (3.75 – 9.00) |  | 10.50 (5.00 – 15.25) |  |
| Treatment group |  |  |  |  |
| Group A | 3.00 (2.00 – 6.50) | 0.473 | 6.00 (5.00 – 9.00) | 0.632 |
| Subgroup A1 | 3.00 (2.00 – 6.00) |  | 6.00 (4.00 – 9.00) |  |
| Subgroup A2 | 3.00 (2.00 – 6.00) |  | 6.00 (5.00 – 8.50) |  |
| Subgroup A3 | 7.00 (4.00 – 9.50) |  | 13.00 (5.00 – 14.50) |  |
| Group B | 3.00 (2.00 – 6.75) |  | 6.00 (4.00 – 9.75) |  |

## References

1. World Health Organization. Coronavirus disease 2019 (COVID-19): situation report—70 2020 [Available from: https://www.who.int/docs/default-source/coronaviruse/situation-reports/20200330-sitrep-70-covid-19.pdf?sfvrsn=7e0fe3f82.

2. Chen N, Zhou M, Dong X, Qu J, Gong F, Han Y, et al. Epidemiological and clinical characteristics of 99 cases of 2019 novel coronavirus pneumonia in Wuhan, China: a descriptive study. The Lancet. 2020;395(10223):507–13.

3. Rothe C, Schunk M, Sothmann P, Bretzel G, Froeschl G, Wallrauch C, et al. Transmission of 2019-nCoV Infection from an Asymptomatic Contact in Germany. N Engl J Med. 2020;382(10):970–1.

4. Bai Y, Yao L, Wei T, Tian F, Jin D, Chen L, et al. Presumed Asymptomatic Carrier Transmission of COVID-19. JAMA. 2020;323(14):1,406 - 1,7.

5. Chinese national health commission and stat administration of traditional Chinese medicine. Diagnosis and treatment protocol for novel corona viruses pneumonia. 2020.

6. Saudi center for disease prevention and control. Quick interim guide to COVID-19 surveillance case definition and disposition. 2020.

7. Centre for Disease Control and Prevention. Information for Clinicians on Investigational Therapeutics for Patients with COVID-19 2020 [updated April 25, 2020. Available from: https://www.cdc.gov/coronavirus/2019-ncov/hcp/therapeutic-options.html.

8. Colson P, Rolain J, Raoult D. Chloroquine for the 2019 novel coronavirus SARSCoV-2. Int J Antimicrob Agents. 2020.

9. Colson P, Rolain JM, Lagier JC, Brouqui P, Raoult D. Chloroquine and hydroxychloroquine as available weapons to fight COVID-19. Int J Antimicrob Agents. 2020;55(4):105932.

10. Liu J, Cao R, Xu M, Wang X, Zhang H, Hu H, et al. Hydroxychloroquine, a less toxic derivative of chloroquine, is effective in inhibiting SARS-CoV-2 infection in vitro. Cell Discov. 2020;6:16.

11. Yao X, Ye F, Zhang M, Cui C, Huang B, Niu P, et al. In Vitro Antiviral Activity and Projection of Optimized Dosing Design of Hydroxychloroquine for the Treatment of Severe Acute Respiratory Syndrome Coronavirus 2 (SARS-CoV-2). Clin Infect Dis. 2020.

12. Wang M, Cao R, Zhang L, Yang X, Liu J, Xu M, et al. Remdesivir and chloroquine effectively inhibit the recently emerged novel coronavirus (2019-nCoV) in vitro. Cell Res. 2020;30(3):269–71.

13. Gautret P, Lagier JC, Parola P, Hoang VT, Meddeb L, Mailhe M, et al. Hydroxychloroquine and azithromycin as a treatment of COVID-19: results of an open-label nonrandomized clinical trial. Int J Antimicrob Agents. 2020: 105949.

14. Chen J, Liu D, Liu L, Liu P, Xu Q, Xia L. A pilot study of hydroxychloroquine in treatment of patients with common coronavirus disease-19 (COVID-19). J of Zjeijang University. 2020;49(1).

15. Ferner RE, Aronson JK. Chloroquine and hydroxychloroquine in covid-19. BMJ. 2020;369:m1432.

16. Gunja N, Roberts D, McCoubrie D, Lamberth P, Jan A, Simes D, et al. Survival after massive hydroxychloroquine overdose. Anaesth Intensive Care 2009;37:130 - 3.

17. Chorin E, Dai M, Shulman E, Wadhwani L, Bar Cohen R, Barbhaiya C, et al. The QT interval in patients with SARS-CoV-2 infection treated with hydroxychloroquine/azithromycin. [[Preprint]]. In press 2020.

18. Makin A, Wendon J, Fitt S, Portmann B, Williams R. Fulminant hepatic failure secondary to hydroxychloroquine. Gut. 1994;35:569 - 70.

19. Murphy M, Carmichael A. Fatal toxic epidermal necrolysis associated with hydroxychloroquine. Clin Exp Dermatol. 2001;26(5):457 - 8.

20. Zhou F, Yu T, Du R, Fan G, Liu Y, Liu Z, et al. Clinical course and risk factors for mortality of adult inpatients with COVID-19 in Wuhan, China: a retrospective cohort study. The Lancet. 2020;395(10229):1054–62.

21. Gautret P, Lagier JC, Parola P, Hoang VT, Meddeb L, Sevestre J, et al. Clinical and microbiological effect of a combination of hydroxychloroquine and azithromycin in 80 COVID-19 patients with at least a six-day follow up: A pilot observational study. Travel Med Infect Dis. 2020: 101663.

22. Saudi Ministry of Health. Protocol for Adults Patients Suspected of/Confirmed with COVID-19 Supportive care and antiviral treatment of suspected or confirmed COVID-19 infection. 2020.

23. Magagnoli J, Narendran S, Pereira F, Cummings T, Hardin JW, Sutton SS, et al. Outcomes of hydroxychloroquine usage in United States veterans hospitalized with Covid-19 [Preprint]. In press 2020.

